# Sex Differences in People with Tourette Syndrome and Persistent Motor or Vocal Tic Disorder in the Tourette Association of America International Consortium for Genetics Database

**DOI:** 10.1101/2024.01.07.24300816

**Authors:** Marisela E. Dy-Hollins, Lori B. Chibnik, Natasha A. Tracy, Lisa Osiecki, Cathy L. Budman, Danielle C. Cath, Marco A. Grados, Robert A. King, Lyon Gholson, Guy A. Rouleau, Paul Sandor, Harvey S. Singer, Nutan Sharma, Carol A. Mathews, Jeremiah Scharf

## Abstract

**Background and Objective:** Tourette Syndrome (TS) and Persistent Motor or Vocal Tic Disorders (PMVT) are more prevalent in males (vs. females). Females with TS may have a delay in diagnosis, and more complex tic features (vs. males). With respect to comorbidities, obsessive-compulsive disorder (OCD) is more prevalent in females; attention-deficit hyperactivity disorder (ADHD) is more prevalent in males. Less is known about sex differences in PMVT. This study analyzes sex differences in outcomes among individuals with TS and PMVT in the Tourette Association of America International Consortium for Genetics dataset (TAAICG).

**Design/Methods:** Data from 2403 individuals (N=2109 TS; N=294 PMVT) from the TAAICG were analyzed to explore the relationship between sex and TS or PMVT outcomes: age at tic onset; age at diagnosis; time-to-diagnosis; tic severity; and comorbidity rates. Regression models were adjusted for age and family relationships to examine the impact of sex on outcomes.

**Results:** Females with TS (25.5% of the sample) had a later age of symptom onset (6.5±2.8 vs. 6.0±2.7; p=0.001), later age at diagnosis (13.3±11.2 vs. 10.7±8.1; p=0.0001), and a longer time-to-diagnosis [3 (1,7) vs. 2 (1,5), p=0.01] than males. The total Yale-Global Tic Severity Scale (YGTSS) was lower in females with TS (28.4±9.1 vs. 30.7±8.7); p<0.0001); OCD was slightly more prevalent in females (55% vs. 48.7%; p=0.01) although OCD severity did not differ by sex; ADHD was more prevalent in males (55.7% vs 38.9%; p<0.001). Females with TS had 0.46 lower odds of being diagnosed with TS (p<0.00001). Females with PMVT (42.9% of the sample) had an earlier age of symptom onset (7.9±3.3 vs. 8.9±3.7; p=0.05). Motor or vocal tic severity (YGTSS) was not significantly different. OCD, but not ADHD, was more prevalent in females (OCD: 41.9% vs. 22.2%; p<0.001: ADHD:16.5% vs 21.0%; p=0.4).

**Conclusion:** Females with TS are less likely to be formally diagnosed and have a later age of symptom onset, later age at diagnosis, longer time-to-diagnosis, higher prevalence of OCD, and lower prevalence of ADHD (vs. males). Females with PMVT have an earlier age of symptom onset, higher prevalence of OCD, but similar ADHD prevalence rates (vs. males). Females with TS and PMVT may be clinically different than males with TS. Future research is needed to understand differences longitudinally in TS and PMVT.

## Introduction

Tic disorders are relatively common neurodevelopmental disorders with prevalence rates of up to 20% in school-age children.^1^ Although the prevalence of tic disorders is lower in females than males (i.e., 4:1 in Tourette Syndrome (TS); 2:1 in chronic/persistent motor or vocal tic disorders [PMVT]),^2–5^ few studies have examined sex differences in natural history and health outcomes among individuals with tic disorders.^6–14^ Individuals with TS have high rates of co-occurring neuropsychiatric conditions, most commonly obsessive-compulsive disorder (OCD) and attention deficit hyperactivity disorder (ADHD).^3^ It has been suggested that females with tic disorders, specifically with TS, may experience delayed diagnosis, worse tic severity in adulthood, and have varying responses to medications compared to males with TS.^6,7,9^ Females with TS have higher rates of OCD compared to males with TS, who in turn have higher rates of ADHD.^10^ Being female is also an independent predictor of worse scores on measures of tic-specific impairment, independent of tic severity, compared to males with tic disorders.^15^ There is less known about sex differences in PMVT.

In this present study, we aimed to examine sex differences among individuals with TS and PMVT using the Tourette Association of America International Consortium for Genetics (TAAICG) dataset in the following outcomes: age of tic symptom onset, age at diagnosis, time-to-diagnosis, tic severity, comorbidity rates of ADHD and OCD, and severity of OCD. We hypothesized that females with TS and PMVT would have worse outcomes compared to males with TS and PMVT.

## Methods

### Study Population

We examined the Tourette Association of America International Consortium for Genetics research dataset of individuals with TS and PMVT and their family members who were recruited for genetics research as part of a study funded by the NIH.^16,17^ These individuals received formal structured diagnostic assessments for the diagnosis of TS or PMVT and associated comorbidities such as ADHD, and OCD, along with validated severity scales (i.e., Yale Global Tic Severity Scale and Yale-Brown Obsessive-Compulsive Scale) as previously described.^10,16,17^ The institutional review board at the Massachusetts General Brigham System approved all aspects of this work. Inclusion and exclusion criteria have been previously reported.^10^

Demographic measures included individuals’ age at interview (years) and biological sex assigned at birth (male, female). Clinical information extracted included individuals’ age of tic symptom onset, age at diagnosis of TS (not available in PMVT individuals), if ever diagnosed with TS (not available in PMVT individuals), best estimate diagnoses of ADHD and/or OCD, and best estimate age of symptom onset of ADHD and/or OCD. Measures used to assess lifetime worst-ever tic-related severity and worst-ever OCD symptom severity were the Yale Global Tic Severity Score (YGTSS),^18^ and the Yale-Brown Obsessive-Compulsive Scale (YBOCS).^19^

### Statistical analysis

For individuals with TS, the primary variables were the age of tic symptom onset, age at diagnosis of TS, time-to-diagnosis of TS, tic severity (individual scores lifetime worst-ever YGTSS Total Tic Score), if ever diagnosed with TS (not available for PMVT). For individuals with PMVT, the primary variables were the age of tic symptom onset, tic severity measured with the YGTSS motor component or the YGTSS vocal component. The age of PMVT diagnosis was not available. For individuals with TS and PMVT, the variables were the best estimate diagnosis of OCD, best estimate age of OCD symptom onset, OCD symptom severity measured with the YBOCS, best estimate diagnosis of ADHD, and best estimate age of ADHD symptom onset.

T-tests were performed for normally distributed continuous outcome variables: age at interview, tic age of symptom onset, TS age of diagnosis, YGTSS tic severity scores, worst ever motor or vocal impairment, OCD age of symptom onset for TS, YBOCS symptom severity scores for TS, and ADHD age of symptom onset for TS. Wilcoxon rank sum tests were performed for non-normally distributed continuous outcome variables: TS time-to-diagnosis, age of OCD symptom onset for PMVT, YBOCS for PMVT, and ADHD age of symptom onset for PMVT. Chi-square tests were performed for categorical outcomes such as if ever diagnosed with TS and best estimate OCD and ADHD diagnosis. We fitted linear and logistic regression models to estimate the association between biological sex and outcomes of interest after adjusting for age at the interview, and clustering for family relationships. Statistical significance was set at the p=0.05 level. Data were analyzed using STATA 17.0 (College Station, TX).

### Sensitivity analysis

To examine the contribution of age and family relationships, a secondary analysis of sex differences was conducted in individuals with TS and PMVT restricted to under 19 years of age. In addition, a secondary analysis was conducted to examine sex differences in the individual components of the YGTSS and lifetime worst-ever motor or vocal impairment.

## Results

There were 2109 individuals with TS and 294 individuals with PMVT in our study sample. The sample consisted of both children recruited for the TS genetic studies, and parents and siblings who were found to be affected at the time of the assessment. Individuals with TS were younger than those with PMVT. 33% of individuals (N=694) with TS were older than age 18 at time of the interview. 357 of these individuals with TS were probands and siblings and 330 were parents. 81.3% of individuals (N=239) with PMVT were older than 18 at the time of the interview. Eleven of these individuals with PMVT were probands and siblings and 216 were parents. The cohort was stratified by males and females with TS and PMVT. Tables 1 and 3 summarize the clinical characteristics of the individuals.

**Table 1.** Sex Differences in Individuals with Tourette Syndrome in the TAAICG Database.

| **Tourette Syndrome (TS)** | **All individuals with TS**<br>**N=2109** | **Female**<br>**N=537 (25.5%)** | **Male**<br>**N=1572 (74.5%)** | **Test statistic** | **p-value** |
| --- | --- | --- | --- | --- | --- |
| Age at Interview (Years), Mean (SD)<br>(N=694/2109, 33% over age 18 year at time of interview) | 19.3 (13.9) | **23.0 (15.1)** | 18.0 (13.2) | -6.8 | **<0.0001** |
| Age at Interview (Years), Median (Q1, Q3) | 13 (10, 23) | **23.0 (15.1)** | 18.0 (13.2) | -6.8 | **<0.0001** |
| **Tic Measures** |  |  |  |  |  |
| Ever Diagnosed with TS | 1485 | **307 (60.8)** | 1148 (77.3) | 52.3 | **<0.001** |
| Age of Symptom Onset (Years), Mean (SD) | 6.2 (2.7) | **6.5 (2.8)** | 6.0 (2.7) | -3.3 | **0.0011** |
| Age of Diagnosis (Years), Mean (SD) | 11.3 (8.9) | **13.3 (11.2)** | 10.7 (8.1) | -3.8 | **0.0001** |
| Time-to-Diagnosis (Years), Median (Q1, Q3) | 3 (1,5) | **3 (1, 7)** | 2 (1, 5) | -2.5 | **0.0111** |
| YGTSS Motor, Mean (SD) (scores >0) | 17.1 (4.5) | **16.4 (4.7)** | 17.4 (4.4) | 4.2 | **<0.0001** |
| YGTSS Vocal, Mean (SD) (scores >0) | 13.3 (5.0) | **12.4 (5.0)** | 13.6 (5.0) | 4.5 | **<0.0001** |
| YGTSS Total, Mean (SD) | 30.2 (8.8) | **28.4 (9.1)** | 30.7 (8.7) | 4.9 | **<0.0001** |
| **OCD Measures** |  |  |  |  |  |
| Best Estimate Diagnosis | 1054 (50.5%) | **291 (55.0%)** | 751 (48.7%) | 6.2 | **0.013** |
| Age of Symptom Onset (Years), Mean (SD) | 7.5 (4.3) | **8.1 (5.5)** | 7.3 (3.8) | -1.9 | **0.06** |
| YBOCS, Mean (SD) (scores >0) | 19.6 (9.3) | 20.3 (9.3) | 19.4 (9.3) | -0.8 | 0.4 |
| **ADHD Measures** |  |  |  |  |  |
| Best Estimate Diagnosis | 1041 (51.4%) | **199 (38.9%)** | 835 (55.7%) | 43.0 | **<0.001** |
| Age Symptom Onset (Years), Mean (SD) | 4.4 (1.7) | 4.6 (1.8) | 4.3 (1.7) | -1.4 | 0.16 |

**Abbreviations:**
TAAICG= Tourette Association International Consortium for Genetics; SD= Standard Deviation;
YGTSS=Yale Global Tic Severity Scale; OCD= Obsessive-Compulsive Disorder; YBOCS= Yale-Brown Obsessive-Compulsive Scale; ADHD= Attention Deficit Hyperactivity Disorder
- bold indicates p <0.05

**Table 2.** Tourette Syndrome: Regression models adjusting for age at interview, and clustering by family relationships

| **Tourette Syndrome (TS) Outcomes** | **Coefficient** | **SE** | **t-value** | **p-value** |
| --- | --- | --- | --- | --- |
| **Age of Symptom Onset (Years)** |  |  |  |  |
| Female | 0.12 | 0.13 | 0.92 | 0.36 |
| Age (Years) | 0.07 | 0.004 | 14.97 | <0.001 |
| **Age of Diagnosis (Years)** |  |  |  |  |
| Female | 0.64 | 0.34 | 1.88 | 0.060 |
| Age (Years) | 0.65 | 0.02 | 29.7 | <0.001 |
| **Time-to-Diagnosis (Years)** |  |  |  |  |
| Female | 0.77 | 0.35 | 2.2 | 0.03 |
| Age (Years) | 0.57 | 0.02 | 26.1 | <0.001 |
| **YGTSS Total** |  |  |  |  |
| Female | -1.67 | 0.48 | -3.5 | <0.001 |
| Age (Years) | -0.14 | 0.01 | -9.5 | <0.001 |
| **TS Outcomes** | **Odds Ratio** | **SE** | **95% CI** | **p-value** |
| **OCD Best Estimate Diagnosis** |  |  |  |  |
| Female | 1.3 | 0.13 | 1.03, 1.6 | 0.02 |
| Age (Years) | 1.0 | 0.003 | 0.99, 1.01 | 0.08 |
| **ADHD Best Estimate Diagnosis** |  |  |  |  |
| Female | 0.6 | 0.004 | 0.96, 0.97 | <0.001 |
| Age (Years) | 0.96 | 0.06 | 0.46, 0.71 | <0.001 |
| **TS + ADHD + OCD** |  |  |  |  |
| Female | 0.89 | 0.10 | 0.70, 1.1 | 0.31 |
| Age (Years) | 0.98 | 0.003 | 0.98, 0.99 | <0.001 |

**Abbreviations:**
YGTSS= Yale Global Tic Severity Scale; OCD= Obsessive-Compulsive Disorder; ADHD= Attention Deficit Hyperactivity Disorder

**Table 3.** Sex Differences in Individuals with Persistent Motor or Vocal Tic Disorders in the

| **Persistent Motor or Vocal Tic Disorder (PMVT)** | **All individuals with PMVT**<br>**N=294** | **Female**<br>**N=168**<br>**(42.9%)** | **Male**<br>**N=126**<br>**(57.1%)** | **Test statistic** | **p-value** |
| --- | --- | --- | --- | --- | --- |
| Age at Interview (Years), Mean (SD)<br>(N=239/294, 81.3% over age 18 at time of interview) | 36.5 (14.8) | 36.6 (14.1) | 36.4 (15.3) | -0.07 | 0.9 |
| Age at Interview (Years), Median (Q1, Q3) | 40 (31, 46) | 39 (33, 45) | 41 (22, 46) | 0.7 | 0.5 |
| **Tic Measures** |  |  |  |  |  |
| Age of Symptom Onset (Years), Mean (SD) | 8.5 (3.5) | **7.9 (3.3)** | 8.9 (3.7) | 2.0 | **0.05** |
| YGTSS Motor, Mean(SD)<br>Scores >0 & vocal=0 | 10.8 (4.5) | 11.2 (4.2) | 10.5 (4.7) | -1.0 | 0.3 |
| YGTSS Vocal, Mean (SD)<br>Scores>0 & motor=0 | 8.0 (3.6) | 8.3 (3.7) | 7.7 (3.5) | -0.5 | 0.6 |
| YGTSS, Vocal, Median (Q1, Q3) | 8 (5, 11) | 8 (5.5, 11) | 8 (5,11) | -0.5 | 0.6 |
| **OCD Measures** |  |  |  |  |  |
| Best Estimate Diagnosis | 89 (30.4%) | **52 (41.9%)** | 37 (22.2%) | 13.1 | **<0.001** |
| Age of Symptom Onset (Years), Median (Q1, Q3) | 9 (6, 13) | 8 (6, 10.5) | 10 (5, 14) | 0.7 | 0.5 |
| YBOCS, Median (Q1, Q3) | 18 (10.5, 21.5) | 15 (10, 18) | 19 (11, 24) | 1.0 | 0.3 |
| **ADHD Measures** |  |  |  |  |  |
| ADHD Best Estimate Diagnosis | 56 (19.3%) | 20 (16.5%) | 35 (21.0%) | 0.9 | 0.4 |
| Age of Symptom Onset (Years), Median (Q1, Q3) | 5 (4, 6) | 5 (5, 6) | 5 (4, 6) | -0.5 | 0.6 |

**TAAICG Dataset\***
\*excludes 2 individuals with age of tic symptom onset > 18 years of age
TAAICG= Tourette Association International Consortium for Genetics;
YGTSS= Yale Global Tic Severity Scale; OCD=Obsessive-Compulsive Disorder; YBOCS= Yale-Brown Obsessive-Compulsive Scale; ADHD= Attention Deficit Hyperactivity Disorder
- bold indicates p <0.05

## Individuals with TS: Comparing females and males (Table 1)

There were 25.5% females (N=537) and 74.5% males (N=1572) with TS in the sample. Of these individuals, 43.3% of the females (N=143) and 56.7% of the males (N=187) were parents.

### Age at time of encounter

Females were older at the time of encounter compared to males (23.0 ± 15.1 years vs. 18.0 ± 13.2 years, t=-6.8; p=<0.0001).

### Ever diagnosed with TS

Females with TS were less likely to have received a diagnosis of TS prior to the research assessment compared to males (60.8% vs 77.3% males, p<0.001). Females with TS had 0.46 times lower odds of ever formally being diagnosed with TS compared to males (p<0.0001).

### Age at of tic symptom onset, age of TS diagnosis, time-to-TS-diagnosis

When compared to males, females also had a later reported age of tic symptom onset (6.5±2.8 years vs. 6.0±2.7 years, t= -3.3 p= 0.001), later mean age of TS diagnosis (13.3±11.2 years vs.10.8±8.1 years, t= -3.8; p=0.001), and a longer time-to-diagnosis of TS by one year (t= -2.5; p=0.01).

### Comorbidities: OCD and ADHD measures

OCD prevalence was higher in females (55%) compared to males (48.7%) (p=0.01). Females had a later age of symptom onset compared to males, although this difference did not reach statistical significance (8.1 ± 5.5 years vs. 7.3 ±3.8 years; t= -0.5; p=0.06). There were no differences observed in mean YBOCS severity scores (20.3±9.3 vs. 19.4±9.3; t= -0.8; p=0.4) between females and males. ADHD prevalence was higher in males (55.7%) compared to females (38.9%) (p<0.001). There was no difference in the mean age of ADHD symptom onset between females and males (4.6±1.8 years vs 4.3±1.7 years; t=-1.4; p=0.16). Age of ADHD diagnosis was not available (Table 1).

### Regression adjusting for age and family relationships *(Table 2)*

After adjusting for age and family relationships, the differences seen with sex and age of symptom onset and age at diagnosis were no longer significant, although the differences between males and females with TS remained significant for time-to-diagnosis and tic severity. Females with TS had 1.3 times higher odds of having OCD compared to males (p=0.02) and 0.6 times lower odds of having ADHD compared to males (p<0.001). There were no differences between males and females with TS and both ADHD and OCD.

## Sensitivity Analyses

### Tic severity and impairment (Supplementary Table 1)

Females with TS had lower tic severity compared to males. The individual motor YGTSS (16.4±4.7 vs 17.4±4.4; t=4.2; p<0.0001) and vocal YGTSS severity scores (12.4±5.0 vs 13.6±5.0; t=4.5; p <0.0001) were lower for females compared to males. The mean lifetime we-YGTSS tic severity was lower in females compared to males (28.4±9.1 vs. 30.7±8.7, t=4.9; p<0.0001).

Females had lower lifetime worst ever phonic tic impairment (2±1.5 vs 2.3±1.5); t=2.1; p=0.03) compared to males. There were no sex differences in lifetime worst ever motor tic impairment.

On the individual YGTSS items, females had lower number (vocal only), intensity (motor and vocal), and complexity (motor and vocal) of tics compared to males. There were no sex differences in number of motor tics. However, females had higher frequency and forcefulness of vocal tics, but lower frequency and forcefulness of motor tics compared to males.

### Restricting age to individuals 18 and under (Supplementary Table 2)

Given that there were parents in the sample, an additional sensitivity analysis was conducted for individuals under 19 years of age to eliminate bias introduced by including parents. As we found in the larger sample, females with TS were less likely to have ever received a formal diagnosis of TS compared to males (73.8% vs 84.7%; p<0.001). Females with TS had 0.5 times lower odds of being diagnosed with TS compared to males (p<0.0001). Females had lower tic severity (individual motor and vocal, total YGTSS) compared to males. The prevalence of ADHD was lower in females (46.4% vs 63.1%; p<0.001). Unlike in the larger sample, there were no sex differences in age of interview, age of tic symptom onset, age of tic diagnosis, and time-to-diagnosis. There were also no sex differences in OCD prevalence.

## Individuals with PMVT: Comparing females and males (Table 3)

A similar strategy was used to analyze sex differences in PMVT. There were 42.9% females (N=126) and 56.7% males (N=168) with PMVT. Most individuals with PMVT were parents of TS-affected children (90.4%). Of these individuals, 46.3% of the females (N=100) and 53.7% of the males (N=116) were parents.

### Age at time of encounter

There were no differences in age at interview in females vs males (36.6±14.1 years vs. 36.4±15.3 years; t=-0.07; p=0.9).

### Age at tic symptom onset

Females reported earlier tic symptom onset than males (7.9±3.3 years vs. 8.9±3.7 years; t=2.0, p=0.05). Age at PMVT diagnosis was not available.

### Comorbidities: OCD and ADHD measures

The prevalence of OCD was higher in females (41.9%) than in males (22.2%) (p<0.001). There were no statistically significant differences in OCD median age of symptom onset [8 (6, 10.5) years vs. 10 (5, 14) years; t=0.7, p=0.5], median YBOCS severity scores [15 (10, 18) vs. 19 (11,24); t= 1.0; p=0.3], ADHD prevalence (16.5% vs 21.0%; p=0.4), or median ADHD age of symptom onset [5 (5, 6) vs. 5 (4, 6); t=-0.5; p=0.6] between females and males.

### Regression adjusting for age and family relationships *(Table 4)*

After adjusting for age and family relationships, the odds of having OCD were 2.4 times higher for females with PMVT compared to males (p=0.001). Females with PMVT had 1.7 times higher odds of having both ADHD And OCD compared to males (0.03). There were no differences seen between males and females with PMVT for ADHD prevalence.

**Table 4.**
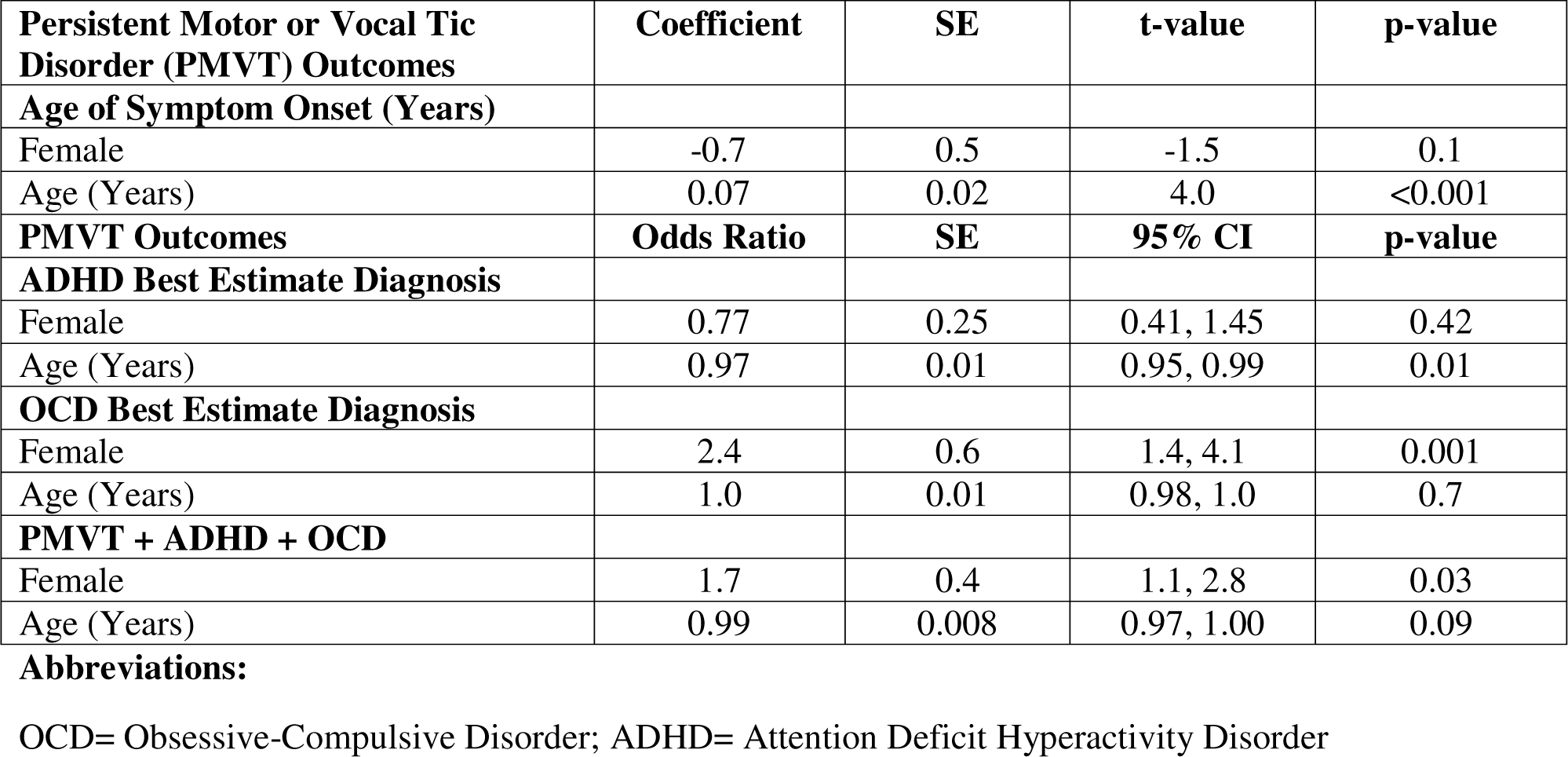
Persistent Motor or Vocal Tic Disorder: Regression models adjusting for sex, age at interview, and clustering by family relationships

| **Persistent Motor or Vocal Tic Disorder (PMVT) Outcomes** | **Coefficient** | **SE** | **t-value** | **p-value** |
| --- | --- | --- | --- | --- |
| **Age of Symptom Onset (Years)** |  |  |  |  |
| Female | -0.7 | 0.5 | -1.5 | 0.1 |
| Age (Years) | 0.07 | 0.02 | 4.0 | <0.001 |
| **PMVT Outcomes** | **Odds Ratio** | **SE** | **95% CI** | **p-value** |
| **ADHD Best Estimate Diagnosis** |  |  |  |  |
| Female | 0.77 | 0.25 | 0.41, 1.45 | 0.42 |
| Age (Years) | 0.97 | 0.01 | 0.95, 0.99 | 0.01 |
| **OCD Best Estimate Diagnosis** |  |  |  |  |
| Female | 2.4 | 0.6 | 1.4, 4.1 | 0.001 |
| Age (Years) | 1.0 | 0.01 | 0.98, 1.0 | 0.7 |
| **PMVT + ADHD + OCD** |  |  |  |  |
| Female | 1.7 | 0.4 | 1.1, 2.8 | 0.03 |
| Age (Years) | 0.99 | 0.008 | 0.97, 1.00 | 0.09 |

**Abbreviations:**
OCD= Obsessive-Compulsive Disorder; ADHD= Attention Deficit Hyperactivity Disorder

## Sensitivity Analyses

### Tic severity and impairment (Supplementary Table 3)

There were no differences in the mean lifetime we-YGTSS motor (11.2±4.2 vs. 10.5±4.7; t=-1.0, p=0.3) or vocal tic severity scores (8.3±3.7 vs. 7.7±3.5; t=-0.5; p=0.6) between females and males. There were no sex differences in lifetime worst-ever tic impairment. There were also no sex differences in the individual YGTSS items.

### Restricting age to individuals 18 and under (Supplementary Table 4)

In the over age 18 group, there were more males (67.3%) with PMVT than females (32.7%). Females with PMVT were younger than males with PMVT (9.9±2.9 vs 12.8±4.2; t=2.9; p=0.005). There were also no sex differences in age of tic symptom onset nor on the individual motor or vocal YGTSS scores. In addition, there were no sex differences in OCD nor ADHD prevalence. Of note, females with PMVT were almost two times more likely to have an OCD diagnosis compared to males, similar to the larger analysis, however, the sample size was small and the differences were not statistically significant.

## Discussion

In this retrospective study, we examined sex differences in clinical outcomes of individuals with TS and PMVT using a large, international clinical dataset from the TAAICG. Consistent with the known epidemiology, the male-to-female ratio of the TS sample was approximately 3:1 (74.5% males and 25.5% females). Females with TS are less likely to have ever received a formal diagnosis of TS (i.e., odds ratio 0.46 times lower in females compared to males). Females with TS were older at time of interview, had a later age of symptom onset, a later age at TS diagnosis, and a longer time-to-diagnosis by one year compared to males with TS. On the individual motor YGTSS items, females had lower frequency, intensity, interference, and complexity compared to males. On the individual vocal YGTSS items, females had lower number, interference, and complexity, but higher frequency and intensity compared to males. In addition, mean lifetime tic severity, as measured by we-YGTSS, was lower in females compared to males, although the differences (mean of 2 points on a 50-point scale) are not likely to be clinically meaningful.

In terms of comorbidities, OCD was more prevalent, and ADHD was less prevalent in females compared to males, which was consistent with what has been reported previously.^10^ Our findings of lower we-YGTSS (and individual motor tic score) in females is in contrast to others who have reported higher tic severity in females on the YGTSS compared to males.^13,14^ However, these differences could be related to methodologic differences in the studies given selection criteria for the cohorts and also age ranges.

For individuals with PMVT, there were 42.9% females and 57.1% males. We found no differences in age at interview when examining sex differences in individuals with PMVT. As noted, the PMVT group was older compared to the TS cohort, and primarily, although not exclusively, consisted of parents of TS-affected individuals. In the PMVT sample, in contrast to the TS sample, females had an earlier age of tic symptom onset compared to males. There were no significant differences found in individual motor or vocal tic severity scores. However, when comparing individual motor and vocal tic severity in individuals with TS and PMVT, individuals with PMVT had both lower motor and vocal YGTSS scores compared to individuals with TS (data not shown). As was seen with the TS sample, OCD was more prevalent among females, and OCD symptom severity (i.e., YBOCS) was not significantly different between males and females. Unlike the TS sample, however, there were no differences in ADHD prevalence nor in age of symptom onset.

The finding that the age of tic symptom onset was earlier in females with PMVT, but later in females with TS than in males could be related to recall bias, as many individuals with PMVT were older compared to individuals with TS in our study. In addition, for the younger participants, the information likely came from their parents as they might be too young or not aware of all their tics, particularly if the tics were mild. It is notable that for both sexes, the age at symptom onset was nearly two years earlier for TS than for PVMT. Similarly, both motor and vocal tic severity were higher in the TS sample than in the PVMT sample, providing support for the suggestion that PMVT may be a milder manifestation of TS.^20^ In the TS, although OCD was slightly more prevalent in females compared to males, but the proportions were similar (55% for females and 48.7% for males). For ADHD, the proportions were 38.9% for females and 55.7% for males. However, for PMVT, the rates of OCD were almost two times higher in females compared to males (41.9% vs 22.2%) and the ADHD rates were similar (16.5% for females and 21.0% for males). The sensitivity analyses demonstrated that females with PMVT were almost two times more likely to have OCD compared to males, however, the sample size was likely too small to detect a significant difference.

This study has several strengths including using well-characterized group of individuals with TS and PMVT and associated comorbid conditions using a best-estimate process. Our study also included validated measures of tics and OCD severity (e.g., we-YGTSS and YBOCS) administered by well trained staff. We also included individuals across the lifespan for both TS and PMVT. Finally, we examined sex differences in a large cohort of individuals with PMVT, which is an area that is understudied with respect to sex differences.

There are limitations to our study. First, the individuals were recruited for genetics research in a study focused on TS. Tic diagnosis for PMVT was not obtained. Individuals with PMVT were older than those with TS; there could be recall bias. Finally, the results of our study may not be generalizable across all patients.

## Conclusion

We demonstrate disparities in diagnosis of TS in females compared to males. Females were less likely to have received a diagnosis of TS compared to males. We also demonstrate sex differences in individuals with TS and PMVT using the TAAICG dataset. Our future research efforts are focused on examining sex differences in larger administrative datasets for both TS and PMVT.

## Data Availability

Data will be shared through collaborative arrangements with qualified investigators.

## Contributions

Marisela E. Dy-Hollins: design/conceptualization of the study; analysis and interpretation of the data; drafting and revising the manuscript for intellectual content

Lori B. Chibnik: design/conceptualization of the study; analysis and interpretation of the data; drafting and revising the manuscript for intellectual content

Natasha A. Tracy: drafting and revising the manuscript for intellectual content

Lisa Osiecki: role in acquisition of the data, revising the manuscript for intellectual content

Nutan Sharma: analysis and interpretation of the data; revising the manuscript for intellectual content

Cathy L. Budman: role in acquisition of the data, design/conceptualization of the study; revising the manuscript for intellectual content

Danielle C. Cath: role in acquisition of the data, design/conceptualization of the study; revising the manuscript for intellectual content

Marco A. Grados: role in acquisition of the data, design/conceptualization of the study; revising the manuscript for intellectual content

Robert A. King: role in acquisition of the data, design/conceptualization of the study; revising the manuscript for intellectual content

Gholson Lyon: role in acquisition of the data, design/conceptualization of the study; revising the manuscript for intellectual content

Guy A. Rouleau: role in acquisition of the data, design/conceptualization of the study; revising the manuscript for intellectual content

Paul Sandor: role in acquisition of the data, design/conceptualization of the study; revising the manuscript for intellectual content

Harvey S. Singer: role in acquisition of the data, design/conceptualization of the study; revising the manuscript for intellectual content

Carol A. Mathews: role in acquisition of the data, design/conceptualization of the study; analysis and interpretation of the data; drafting and revising the manuscript for intellectual content

Jeremiah M. Scharf: role in acquisition of the data, design/conceptualization of the study; analysis and interpretation of the data; drafting and revising the manuscript for intellectual content

**Search Terms:** sex differences, Tourette Syndrome, Persistent Motor or Vocal Tics (Chronic Tics), disparities

**Data Sharing:** Data will be shared through collaborative arrangements with qualified investigators.

**Study Funding:** This study was funded by K12NS098482, R01 NS102371, and R01 NS105746. The content is solely the responsibility of the authors and does not necessarily represent the official views of the National Institutes of Health.

## Notification of any redundant or duplicate publication

None

## Acknowledgments

This study was funded by the Tourette Association of America and NIH K12NS098482, R01 NS102371, and R01 NS105746. We are grateful to the individuals with TS and their families that participated in the study. We also acknowledge members of the Tourette Association of America International Consortium for Genetics.

---

**Supplementary Table 1.** Sex Differences in Individuals with Tourette Syndrome in the TAAICG Database-Individual Motor and Vocal YGTSS Scores

| **Tourette Syndrome (TS)** | **All individuals with TS**<br>**N=2109** | **Female**<br>**N=537 (25.5%)** | **Male**<br>**N=1572 (74.5%)** | **Test statistic** | **p-value** |
| --- | --- | --- | --- | --- | --- |
| **Worst Ever Impairment**<br>(Range 0-5) |  |  |  |  |  |
| Motor, Mean (SD) | 2.87 (1.4) | 2.7 (1.5) | 2.9 (1.4) | 1.6 | 0.1 |
| Vocal, Mean (SD) | 2.2 (1.5) | **2 (1.5)** | 2.3 (1.5) | 2.1 | **0.03** |
| **Number** (Range 0-5) |  |  |  |  |  |
| Motor, Mean (SD) | 3.36 (1.1) | 3.3 (1.1) | 3.4 (.1) | 1.4 | 0.16 |
| Vocal, Mean (SD) | 2.2 (0.83) | **2.1 (0.9)** | 2.2 (0.8) | 2.4 | **0.02** |
| **Frequency** (Range 0-5) |  |  |  |  |  |
| Motor, Mean (SD) | 3.9 (1.2) | **3.7 (1.3)** | 3.9 (1.2) | 3.6 | **0.0004** |
| Vocal, Mean (SD) | 3.3 (1.4) | **3.4 (1.4)** | 3.2 (1.5) | 3.2 | **0.0015** |
| **Intensity** (Range 0-5) |  |  |  |  |  |
| Motor, Mean (SD) | 3.5 (1.2) | **3.2 (1.3)** | 3.6 (1.2) | 5.6 | **<0.0001** |
| Vocal, Mean (SD) | 2.9 (1.4) | **3.1 (1.4)** | 2.7 (1.4) | 4.7 | **<0.0001** |
| **Interference** (Range 0-5) |  |  |  |  |  |
| Motor, Mean (SD) | 2.7 (1.5) | **2.6 (1.6)** | 2.8 (1.5) | 3.2 | **0.0013** |
| Vocal, Mean (SD) | 2.3 (1.5) | **2.1 (1.5)** | 2.3 (1.5) | 3.6 | **0.0003** |
| **Complexity** (Range 0-5) |  |  |  |  |  |
| Motor, Mean (SD) | 3.6 (1.4) | **3.4 (1.4)** | 3.7 (1.3) | 3.4 | **0.0007** |
| Vocal, Mean (SD) | 2.3 (2.0) | **1.97 (1.97)** | 2.4 (2.0) | 4.06 | **0.0001** |

- bold indicates p <0.05

**Supplementary Table 2.** Sex Differences in Individuals with Tourette Syndrome in the TAAICG Database-Individuals 18 and under

| **Tourette Syndrome (TS)** | **All Individuals with TS**<br>**N=1415** | **Female**<br>**N=291 (20.6%)** | **Male**<br>**N=1124 (79.4%)** | **Test statistic** | **p-value** |
| --- | --- | --- | --- | --- | --- |
| Age at Interview (Years), Mean (SD) | 11.4 (3.3) | 11.6 (3.3) | 11.4 (3.2) | -1.2 | 0.2 |
| **Tic Measures** |  |  |  |  |  |
| Ever Diagnosed with TS | 1141 | **208 (73.8)** | 933 (84.7) | 18.8 | **<0.001** |
| Age of Symptom Onset (Years), Mean (SD) | 5.6 (2.4) | 5.8 (2.4) | 5.5 (2.4) | -1.5 | 0.1 |
| Age of Diagnosis (Years), Mean (SD) | 8.25 (2.4) | 8.3 (2.9) | 8.3 (2.5) | -0.0045 | 0.99 |
| Time-to-Diagnosis (Years), Median (Q1, Q3) |  | 2 (1,4) | 2 (1,4) | 0.35 | 0.7 |
| YGTSS Motor, Mean (SD) (scores >0) | 17.6 (4.3) | **16.9 (4.8)** | 17.7 (4.2) | 2.7 | **0.008** |
| YGTSS Vocal, Mean (SD) (scores >0) | 13.7 (5.0) | **12.6 (5.2)** | 14.0 (5.0) | 3.9 | **0.0001** |
| YGTSS Total, Mean (SD) | 31.2 (8.2) | **29.5 (8.7)** | 31.7 (7.96) | 3.7 | **0.0002** |
| **OCD Measures** |  |  |  |  |  |
| Best Estimate Diagnosis, | 674 (48.3%) | 143 (50.0%) | 531 (47.8%) | 0.4 | 0.5 |
| Age of Symptom Onset (Years), Mean (SD) | 6.3 (2.8) | 6.4 (2.8 ) | 6.3 (2.8) | -0.2 | 0.8 |
| YBOCS, Mean (SD) (scores >0) | 19.4 (9.2) | 18.3 (8.3) | 19.6 (9.4) | 0.97 | 0.3 |
| **ADHD Measures** |  |  |  |  |  |
| Best Estimate Diagnosis | 819 (59.7%) | **130 (46.4%)** | 689 (63.1%) | 25.8 | **<0.001** |
| Age Symptom Onset (Years), Mean (SD) | 4.3 (1.7) | 4.4 (1.8) | 4.2 (1.6) | -0.8 | 0.4 |

**Abbreviations:**
TAAICG=Tourette Association International Consortium for Genetics; SD= Standard Deviation;
YGTSS= Yale Global Tic Severity Scale; OCD= Obsessive-Compulsive Disorder; YBOCS= Yale-Brown Obsessive-Compulsive Scale; ADHD= Attention Deficit Hyperactivity Disorder
- bold indicates p <0.05

**Supplementary Table 3.** Sex Differences in Individuals with Persistent Motor or Vocal Tic Disorders in the TAAICG Database-Individual Motor and Vocal YGTSS Scores

| **Persistent Motor or Vocal Tic Disorder (PMVT)** | **All individuals with PMVT**<br>**N=294** | **Female**<br>**N=168 (42.9%)** | **Male**<br>**N=126**<br>**(57.1%)** | **Test statistic** | **p-value** |
| --- | --- | --- | --- | --- | --- |
| **Worst Ever Impairment**<br>(Range 0-5) |  |  |  |  |  |
| Motor, Mean (SD) | 1.0 (1.3) | 0.7 (0.9) | 1.2 (1.4) | 1.8 | 0.08 |
| Vocal, Mean (SD) | 0.5 (1.0) | 0.4 (1.0) | 0.5 (0.9) | 0.5 | 0.6 |
| **Number** (Range 0-5) |  |  |  |  |  |
| Motor, Mean (SD) | 2.0 (1.1) | 2 (1.1) | 2 (1.1) | 0.05 | 0.96 |
| Vocal, Mean (SD) | 0.7 (0.8) | 0.7 (0.80) | 0.7 (0.9) | 0.7 | 0.5 |
| **Frequency** (Range 0-5) |  |  |  |  |  |
| Motor, Mean (SD) | 2.2 (1.7) | 2.3 (1.6) | 2.2 (1.7) | -0.2 | 0.9 |
| Vocal, Mean (SD) | 0.8 (1.4) | 0.8 (1.5) | 0.8 (1.4) | 0.3 | 0.8 |
| **Intensity** (Range 0-5) |  |  |  |  |  |
| Motor, Mean (SD) | 2.2 (1.7) | 2.3 (1.6) | 2.2 (1.7) | -0.2 | 0.9 |
| Vocal, Mean (SD) | 0.8 (1.4) | 0.8 (1.5) | 0.8 (1.4) | 0.3 | 0.8 |
| **Interference** (Range 0-5) |  |  |  |  |  |
| Motor, Mean (SD) | 1.2 (1.1) | 1.2 (1.0) | 1.3 (1.1) | 0.4 | 0.7 |
| Vocal, Mean (SD) | 0.5 (0.8) | 0.5 (0.9) | 0.5 (0.8) | -0.2 | 0.8 |
| **Complexity** (Range 0-5) |  |  |  |  |  |
| Motor, Mean (SD) | 2.1 (1.7) | 2.2 (1.8) | 1.9 (1.6) | -1.3 | 0.2 |
| Vocal, Mean (SD) | 0.6 (1.3) | 0.5 (1.3) | 0.6 (1.3) | 0.7 | 0.5 |

**Supplementary Table 4.** Sex Differences in Individuals with Persistent Motor or Vocal Tic Disorder in the TAAICG Database-Individuals 18 and under

| **Persistent Motor or Vocal Tic Disorder (PMVT)** | **All Individuals with PMVT**<br>**N=55** | **Female**<br>**N=18 (32.7%)** | **Male**<br>**N=37 (67.3%)** | **Test statistic** | **p-value** |
| --- | --- | --- | --- | --- | --- |
| Age at Interview (Years), Mean (SD) | 1.9 (4.0) | 9.9 (2.9) | 12.8 (4.2) | 2.9 | 0.005 |
| **Tic Measures** |  |  |  |  |  |
| Age of Symptom Onset (Years), Median (Q1, Q3) | 5.5 (4, 8) | 5 (4, 6.5) | 6 (4, 8) | 0.8 | 0.4 |
| YGTSS Motor, Mean (SD) (scores >0) | 10.6 (7.5) | 9.3 (8.0) | 1.3 (7.2) | 0.8 | 0.4 |
| YGTSS Vocal, Mean (SD) (scores >0) | 2.1 (4.5) | 2.7 (5.0) | 1.8(4.3) | -0.6 | 0.5 |
| **OCD Measures** |  |  |  |  |  |
| Best Estimate Diagnosis | 15 (27.8%) | 7 (38.9%) | 8 (22.2%) | 1.7 | 0.2 |
| Age of Symptom Onset (Years), Mean (SD) |  | N=6 | N=2 | - | - |
| YBOCS, Mean (SD) (scores >0) |  | N=1 | N=2 | - | - |
| **ADHD Measures** |  |  |  |  |  |
| Best Estimate Diagnosis | 3 (21.4%) | N=1 | N=2 | - | - |
| Age Symptom Onset (Years), Mean (SD) |  |  |  | - | - |

**Abbreviations:**
TAAICG=Tourette Association International Consortium for Genetics; SD= Standard Deviation;
YGTSS=Yale Global Tic Severity Scale; OCD= Obsessive-Compulsive Disorder; YBOCS= Yale-Brown Obsessive-Compulsive Scale; ADHD= Attention Deficit Hyperactivity Disorder

